# The social determinants of maternal and congenital syphilis at the Colombia-Venezuela border: A qualitative study of twenty mothers of newborns with congenital syphilis

**DOI:** 10.1101/2024.08.30.24312781

**Authors:** Doris Parada, Andrea Wirtz, Rafael Olarte, Magaly Pedraza, Bert Hoffmann, Merike Blofield

## Abstract

Humanitarian crises and resulting out-migration have created contexts in which treatable diseases such as syphilis have expanded in prevalence. Untreated syphilis can have potentially irreversible and devastating consequences, especially for infants born with congenital syphilis. Our study aimed to understand the experiences of postpartum mothers whose newborns have been diagnosed with congenital syphilis and explore the social determinants of maternal and congenital syphilis. The setting of our study is the main public hospital in Cúcuta, Colombia, at the border with Venezuela, which provides emergency care to everyone regardless of documentation and thus receives a high share of Venezuelan migrants. We conducted twenty in-depth interviews with women who had their newborns hospitalized with a diagnosis of congenital syphilis. Sampling was conducted purposively at the hospital, between March and June 2023. Study participants were mostly Venezuelan migrants and Colombian returnees, from their teens to their forties. We used a grounded theory technique to conduct thematic analysis. Four major themes emerged: 1) experiencing a pregnancy in the context of a lack of resources, violence and ignorance; 2) guilt with and reinfection of syphilis; 3) challenges and limitations in accessing health care; and 4) limited support networks and machismo. Reported challenges were intertwined with the high costs of health care in the country of origin, the lack of knowledge of sexually transmitted infections, the limited public health education targeting this population group, and the absence of Colombian public policies that promote care for the non-regularized migrant population.

## Introduction

The global burden of syphilis has expanded over the last three decades. A 2019 Global Burden of Disease study reported that the age standardized incidence rate of syphilis increased from 160.03/100,000 persons (95% UI: 120.66–208.1) in 1990 to 178.48/100,000 persons (95% UI: 134.94–232.34) in 2019 [1]. In Latin America, the Pan American Health Organization (PAHO) estimated the prevalence of syphilis in men of 0.91%, 0.92% in women (0.92%), and 0.86% among pregnant women in 2016, relatively high compared to other regions [2]. Within the region, the age standardized incidence rate was highest in the Andean region, inclusive of Colombia, Peru, Ecuador, and Bolivia.

Congenital syphilis, that is, the transmission of syphilis via the placenta from mother to fetus that can lead to still birth, miscarriage, and neurological conditions in infants, has increased along with increases in the global burden of syphilis infection [3]. The risk of vertical transplacental transmission due to untreated maternal disease is associated with the stage of maternal syphilis, with 70% to 100% of cases occurring in early stages [4] and leads to fetal or neonatal complications as has been widely documented [5–7].

As with adult syphilis, the incidence of congenital syphilis in the Americas has doubled between 2009 and 2020, from 0.3 to 0.61 per 1000 live births (excluding Brazil) [2]. In Colombia, the rate is significantly higher, increasing from 1.80 per 1000 live births (LB) + stillbirths in 2019 to 2.4 per 1000 LBW + stillbirths in 2023 [8,9]. The Norte de Santander region, bordering Venezuela and the site of the current investigation, presented an incidence rate of 5.7 per 1000 live births in 2022, which was more than double the national rate [8].

While multiple studies have documented the increased prevalence of syphilis among pregnant women and congenital syphilis among vulnerable populations, less is known about the experiences of displaced populations. However, recent and rapid migration trends in Latin America underscore a need to better understand the health conditions for displaced populations, such as Venezuelan migrants and refugees and displaced Colombians.

As a result of a decade-long economic and political crisis, an estimated 7.7 million Venezuelans have been displaced from Venezuela as of November 2023; more than 6.54 million are estimated to have remained in the Latin American and Caribbean region, of whom 2.88 million, or about 44%, are residing in Colombia [10]. Further, Colombia hosts one of the largest populations of internally displaced people, with over 6.8 million people estimated to be internally displaced as of the end of 2022 and more than half a million Colombians who had returned after previously seeking asylum in Venezuela [11]. The causes and contexts of displacement create social vulnerabilities that place people at increased risk for disease, including syphilis, and also likely challenge access to prevention and treatment [12]. Indeed, the population prevalence of syphilis infection among Venezuelan migrants residing in urban settings of Colombia in 2021-22 was estimated at 5.1% (95% CI: 4.6–5.6) and as high as 9.0% (95% CI 5.4–14.6) among pregnant women [13].

The dire conditions in Venezuela have increased the risks of contracting syphilis while decreasing access to treatment [14, 15]. This has motivated many people, especially pregnant women, to cross the border into Colombia to access emergency health care [15]. In addition, many women who already reside in Colombia have problems accessing non-emergency care. Although there is a path for migrants to regularize their status and be registered for the Colombian public health system, many are still not eligible and even among those who are, many have not done so [16]. Prenatal healthcare is not considered an emergency and therefore undocumented migrants do not have routine access to it in public healthcare, although they may have fragmented access through international and local humanitarian programs. (Bogotá, the capital city, has a program that grants all pregnant women access to prenatal care) [17].

As a result, many pregnant women enter the Colombian health system with undiagnosed and untreated syphilis. The Colombian Ministry of Health reports higher prevalence rates among pregnant migrants, who are often diagnosed with syphilis late, even up to the third trimester or during the puerperium [14], which limits adequate treatment and increases the risk of vertical transmission and complications for the mother-child pair [14].

Treatment in both Colombia and in Venezuela follows an adaptation of US CDC guidelines, which recommend benzathine penicillin at a dose of 2,400,000 IU applied intramuscularly at the time of a positive rapid treponemal test. Treatment then continues with the management according to the stage of syphilis in which the pregnant woman is [18, 19].

We aimed to analyze the lived experiences of mothers with newborns with congenital syphilis in a border city of Colombia and, specifically, explore the potential social determinants of health, structural and intermediate, that led to this situation. The main objective is to understand the experiences of mothers with newborns diagnosed with congenital syphilis and treated at the public hospital during 2023.

## Data and methods

We used a qualitative, phenomenological approach, which seeks to deepen the lived experiences, to understand the participant, and to know him/her from the existential experience within his/her daily life, in what Husserl calls the world of life [20]. Thus, we focused on a given situation and group, which allows for the analysis of dynamics in an intersubjective world that is being constructed in the midst of interactions, languages and contexts, in which diverse meanings and motivations emerge [21]. To ensure methodological rigor, we considered the criteria of credibility, auditability and transferability [22] and met all criteria outlined in the COREQ checklist for qualitative research [23].

### Setting and Participants

We conducted the study in Cúcuta, a border city in the province of Norte de Santander, Colombia. Participants were sampled from the main public hospital, the Erasmo Meoz University Hospital (HUEM), that provides medical coverage for most of the population in vulnerable condition of in Cúcuta.

The inclusion criteria were as follows: 1) women with delivery care at HUEM who had newborns presenting a diagnosis of confirmed congenital syphilis; or 2) women in immediate or mid-term postpartum admitted to HUEM with their newborn for treatment for confirmed congenital syphilis; and 3) women who consented to participate in the research. The criteria allowed us to obtain a homogeneous purposive sample, reducing bias. The HUEM epidemiological surveillance office supported sampling by identifying eligible mothers of newborns with confirmed or probable diagnosis of congenital syphilis, hospitalized for treatment or in the puerperium room with their mothers. We estimated that 20 interviews would be needed to achieve thematic saturation, though resource constraints prevented us from sampling to achieve saturation within key subgroups (i.e., Colombian returnees).

### Procedure and ethical considerations

The semi-structured interviews were conducted by a nurse and psychologist who both had extensive experience in qualitative research. The interviews were conducted in Spanish, in a private space and lasted approximately 40 minutes. They were recorded with the participant’s consent.

The project was approved by the HUEM ethics committee (#12-2023, March 7). Once eligible candidates were identified, the nurse and psychologist working on the project approached the mothers, explained the objective of the research project, and ensured written informed consent from each mother for participation in the study, guaranteeing anonymity and confidentiality. All of the above was based on current national and international regulations for the development of studies with human subjects.

### Research tools

We used a semi-structured instrument to guide the interview. The interview guide contained dimensions covering the Social Determinants of Health (SDH). [interview guide as appendix.] The interview guide included questions on structural determinants such as the socioeconomic and political context, access to health, education and work, as well as behavioral and biological factors, living conditions and psychosocial factors.

These questions in turn allowed for the emergence of new questions and an open process of free and flexible interaction between interviewers and participants [24]. This facilitated the expression and open narratives about the experiences of the mothers during and leading up to the pregnancy. The start date of the study was 01-04-2023 and the end date was 16-06-2023. A total of 20 interviews were conducted, achieving theoretical saturation of the data, focused on uncovering the experiences of mothers with infants with congenital syphilis.

### Analysis of the information

Recorded interviews were transcribed and imported into KoBoCollect® (KoBo Inc, Cambridge MA) for analysis. We used grounded theory as our method of analysis [25], systematically implementing open, axial, and selective coding. In open coding, DP reread the interviews line by line, identifying significant findings. As a second step in the analytic process, DP conducted axial coding in which relationships between codes (subcategories) were identified. Finally, DP conducted selective coding to identify emerging categories and central themes related to the experiences of the participating mothers. MB reviewed the coding and analysis. All analyses were conducted in the original Spanish language; final exemplary quotes were translated into English for this manuscript.

## Results

The final study sample included 20 postpartum women diagnosed with gestational syphilis and live newborns diagnosed with congenital syphilis, treated at HUEM, between March and June 2023. Participants ranged in age from 17 to 41 years. The majority were Venezuelan migrant women (n=18, 90%). Fifteen (75%) of the Venezuelan participants had an irregular migratory status. Over half of participants had completed primary education, and 14 reported currently living with a partner. Only 3 of 20 participants were registered in the Colombian public health system, and the majority (n=18, 90%) had an income below the poverty line. Fifteen participants (75%) reported engaging in informal remunerated work. Table 1 outlines the participants’ sociodemographic characteristics.

**Table 1.** Sociodemographic characteristics of participants (N=20)

| Characteristics | N | % |
| --- | --- | --- |
| **Age:** |  |  |
| Teens -28 years | 15 |  |
| 29 years- forties | 5 |  |
| **Venezuelan migrants** | 18 |  |
| **Colombian returnees** | 2 |  |
| **Migration status of Venezuelans** |  |  |
| Irregular | 15 |  |
| Regular | 3 |  |
| **Civil status** |  |  |
| Single | 6 |  |
| Married | 1 |  |
| Cohabiting | 13 |  |
| **Educational level** |  |  |
| Primary school | 11 |  |
| Secondary school | 8 |  |
| University | 1 |  |
| **Number of children** |  |  |
| 1 | 5 |  |
| 2 | 7 |  |
| 3 | 5 |  |
| Over 3 | 3 |  |
| **Engage in paid work** |  |  |
| Yes | 15 |  |
| No | 5 |  |
| **Average monthly income** |  |  |
| USD 6 -14** | 4 |  |
| USD 50 -120 | 4 |  |
| USD 140 - 200 | 5 |  |
| Over USD 240 | 2 |  |
| **Affiliation in Colombian public healthcare** | 3 |  |
| No | 17 |  |

\*\* Colombian extreme poverty level

In terms of reproductive health, participants reported that their first sexual intercourse occurred between the ages of 12 and 18 years and the number of sexual partners in the last year ranged between 1 and 4. Most of the women in the study (n=15) became mothers during adolescence, between 14 and 19 years of age.

Our qualitative analysis identified four main central themes (Table 2), which allowed us to understand the experiences of mothers with babies treated for congenital syphilis at the Erasmo Meoz University Hospital in Cúcuta.

**Table 2.** Central themes and subthemes of experiences of mothers with infants diagnosed with congenital syphilis.

| **Categorías** | **Subcategorías** |
| --- | --- |
| **Experiencing pregnancy amidst migration, lack of resources, and violence** | Irregular migration status<br>Viewing migration as a chance to live with dignity<br>Making miracles with the money that comes in.<br>Overburdened with double shifts with children<br>Living in uncertainty and material insecurity<br>Building a social position amidst vulnerabilities |
| **Lack of knowledge of syphilis leading to acquisition and guilt for diagnosis of mother and child** | "I had never heard of it" (Syphilis)<br>Lack of adherence to treatment for fear of its effects<br>Seeking treatment late due to lack of knowledge<br>Guilt for the child's condition |
| **Challenges and limitations in access to healthcare** | Lack of opportunity to enroll in the health care system |
|  | Inability to afford health services in country of origin.<br>Undignified treatment in country of origin and search for new access to health care<br>Late diagnosis and inadequate number of prenatal visits<br>Colombia as a lifeline for access to health services |
| **The scarcity of support networks and machismo** | Weak state-provided assistance<br>Masculinities that question the safety of treatment<br>Negative emotions towards partners that pave the way for reinfection |

### Theme: “Experiencing pregnancy amidst migration, lack of resources, and violence”

In the case of the 18 Venezuelan women, eight reported that they had moved to Colombia for reasons such as social violence, poor family relationships or, on the contrary, because they have family in Colombia, which provides them with opportunities. The remaining group of migrants (10) report that the main reason for leaving their country was economic, as they perceive job opportunities in Colombia that they could not find in Venezuela even when their current income, for the most part, did not reach the minimum wage, reported that migrating to Colombia it allowed them to subsist in a more dignified manner than when in Venezuela. In general, participants reported that hunger and continuous shortages were constant during their pregnancy.

> *There came a time when I ate bread with water, I did not have enough to eat, because when I started the pregnancy I was not with him, so I recycled and I could not afford anything, one day I ran out of gas … because I could not pay the utilities … now sometimes I could buy bread, I lasted about a month eating bread with water (P17, in her thirties)*.

In the transition to obtaining a stable place to live and work in Colombia, some participants reported that they and their families had to sleep on the streets on several occasions, as well as stay in temporary accommodation where they paid around $6 USD a day. While there are places of refuge established by various non-governmental organizations (NGOs) in Colombia, these women did not have access to them due to the high demand for the service, or because they were unaware of their existence.

> *In the street for up to fifteen days we slept (P10, in her thirties)*.
>
> *Yes, when I arrived when I had nowhere to go, I had to stay in La Parada (border municipality), I was cold and hungry … Yes, I slept in the street… I walked for eight days, I was about three months pregnant, with my [7-12 year old child] (P14, in her twenties).*

Regardless of the situations they endured, women reported suffering extreme poverty in Venezuela that motivated them to migrate and look for a way to provide for their children and families. In some cases, they decided to leave their children in their country of origin to look for a job in Colombia with the possibility of sending remittances, thus solving the problem of food insecurity.

> *Sometimes we had nothing to eat … we lasted three days eating only yucca, then I started to think … I saw people coming and I came. I got a job and then I went to look for the girls (P17, in her thirties)*.

In Colombia, basic needs such as food and housing are better covered. However, participants reported that it was difficult to prioritize healthcare and antenatal care, given that they must focus on meeting the needs of their family.

> *At least for my children, I have them studying, whatever I can and they need and if I can ask the employer, I send to them (they are in Venezuela) to cover their needs, I have been able to buy things for the household … and yes, yes, I have seen improvement, when I was in Venezuela, I had nothing (P2, in her twenties)*.

However, the absence of regularization of legal status complicated these efforts. Of the 18 Venezuelans, only three had regular legal status in Colombia; two held Temporary Protected Status (PPT), which is valid for a 10 year -stay in Colombia, and one had a safe-conduct visa. The women with PPT had legal access to the non-contributory public health system, were eligible to apply for formal employment and potentially be affiliated in contributory social security and health insurance. Of the two Colombian returnees, one did not have documentation but was able to access health services as a Colombian citizen. The remaining group (15) were in an irregular situation, of which five did not have any type of document; nine only had a Venezuelan identity card and one had a Venezuelan Migrant Registration Number, which is a step in the process to access the PPT but does not yet confer the benefits of a regular migratory status. Not having such documentation prevented them from accessing health centers.

> *In Agua Clara (rural area of Cúcuta) … because I did not have the [ID card]*
>
> *so I could not get check-ups there (P11, in her twenties)*.

Further, women were forced to enter Colombia through illegal entry points and unsafe routes.

> *Here … because of the issues do you know how difficult it is? Not having documents, not being able to get a good job, not having a legal document (P14, in her twenties)*.
>
> *I went through the trail and I don’t have an ID card, because I have problems with the document (P2, in her twenties)*.

Twelve Venezuelan women reported pendular migration (moving back and forth) between Colombia and Venezuela. All had been residing in Colombia for between one and twelve months but without regularized legal status.

In terms of income, of the 20 women participating in the study, 15 reported being employed informally as cooks, seamstresses, manicurists, street vendors, pedestrian transporters of goods on the border bridges, or in on-line work.

Working hours of the women were between 4 and 13 hours a day. Their monthly income was diverse, ranging from $6 to $240 dollars a month, except for one participant who earned more.

> *as a [xxx]… I was able to treat my child … because [they] got meningitis and had a valve in [their] head and with that I could buy milk and all the things that ECOOPSOS ([the Colombian public health system]) would not give me (P11, in her twenties)*.

### Theme: “ Lack of knowledge of syphilis leading to acquisition and guilt for diagnosis of mother and child”

The low educational attainment among most of the mothers as well as the minimal sexual and reproductive health education resulted in a lack of awareness of syphilis and was perceived to have resulted in the acquisition of syphilis.

> *I’ll be honest with you: nothing, I don’t know anything about syphilis (P1, migrant, in her teens); it’s one’s fault (congenital syphilis) for not knowing anything (P4, in her teens)*.

Participants reported feeling guilty for not having prevented vertical transmission to their children, as in the following:

> *When I had the baby, the doctor told me that I had tested positive. Yes, it was very hard for me, because if I had known … my baby would not be going through all this, I feel very sad and I know that it is my fault that my baby has to receive all these treatments (P9, in her twenties)*.

Even with a positive test result in their hands, many did not understand how to interpret it. In one of the cases, a participant reported that she had not been informed of her diagnosis by the doctor treating her pregnancy, and learned of her diagnosis only after being seen by another gynecologist:

> *The first doctor evaluated me and did not tell me anything, he only sent me a vaginal cream, when I came here because I was in a lot of pain and the doctor who saw me asked me if I had already seen the tests and I told him that I had been checked, but that he had not told me anything, and he [the attending doctor] told me that they were going to do the tests again and I came back positive … “How strange that this doctor had not told you anything, that you had syphilis”, the doctor told me…, and I told him: “I have syphilis?! no, he didn’t tell me anything”. He asked me if I had ever had injections and I said never and then he gave me the three injections (P10, in her thirties)*.

Participants reported knowing very little if anything about syphilis before being diagnosed with it. For example, some referred to transmission/acquisition risks related to sitting on a toilet or through saliva. Others expressed a belief that it leads to HIV.

Regarding treatment, they believe that it does not disappear completely, but leaves after-effects. Only in the case related to protective measures to avoid acquisition, just over half of participants (11) referred to the need to use condoms with their sexual partners. Participants reported that they had acquired the little information they had predominantly from school and parents, then friends and relatives, and finally the internet.

Sixteen of the mothers reported receiving information about the importance of diagnosing and treating partners to prevent reinfection while they were in the health institutions where they were tested and diagnosed with syphilis. The four mothers who claim not to have received this information were treated in Venezuelan institutions.

> *The first time I was diagnosed, they did not tell me that my partner should also receive the doses (P9, in her twenties)*.

The above quote illustrates the context of a vulnerable population and lack of information and care received in Venezuela.

### Theme: “between challenges and limitations in accessing health care”

Venezuelan participants described difficulty in accessing healthcare in Venezuela, where public entities lack human and material resources. Often, participants reported feeling forced to resort to private care where they had to pay for prenatal check-ups and treatment. In some cases, this made such care unattainable.

> *… I received the first dose, they sent me here to Cúcuta, to have some tests, but I did not have the money to come here… There it was all for pay, each echo and consultation cost me 10 dollars, but I did not have the 9 prenatal visits, I had them every two months, about 5 or so, so I was able to have them done. The first time I paid for the exams, but later, as there was a hospital, I had them done for free, some exams they did and others one had to pay for (P1, in her teens)*.

*In Venezuela, nothing! Here in Colombia, as soon as I showed the exams to the doctor, they started to give me the vaccines (benzathine penicillin); (P16, in her thirties)*.

In Venezuela, participants referred to dehumanized treatment and obstetric violence, which included pejorative words and lack of provision of care required by the mother and child.

> *… in Venezuela they treated me very badly; I am talking about my first child; very badly, I had to give birth on my own, because they put me… and they passed out magazines and then they treated me when they saw that I already had the baby on the stretcher… The check-ups were good, because I paid for them (P13, in her twenties)*.

In Colombia, depending on their migration status, the woman accessed healthcare in different ways. Three had access to the public healthcare system, which allowed them access to prenatal public health services. Access to delivery care, which was open to all regardless of immigration status, was reportedly easy, the treatment more humane and egalitarian, and participants’ stories demonstrated the emotion they felt for the welcome they received in these health facilities.

> *We have been treated very well, thank God; the organizations are also very attentive, the kits help us a lot because we have nothing here. (P9, in her twenties)*.

Some participants referred to a lack of access to prenatal visits, or to the difficulties in access when they do not have the proper legal documents. However, they report having found assistance from NGOs and in some cases in public health services, obtaining care and treatment according to their needs.

The number of syphilis tests performed during pregnancy was between 1 and 4. Some mothers (5) were diagnosed between the 16th and 21st weeks of pregnancy, which made it possible to diagnose and treat it in a timely manner. Five of the mothers were diagnosed only after the 36th week, even in the 39th week, timing of which reduces the ability to limit infectious transmission to the fetus or neonate.

Seven of the women reported not having known their seropositivity for syphilis at some point during pregnancy. Within this group, five reported that they only learned of the diagnosis of congenital syphilis and thus of gestational syphilis at birth, from the serology test performed on newborns, and which then implied the hospitalization of the child and the beginning of the treatment process and conversations with the couple to cut cycles of reinfection.

**Table 4.** Number of prenatal checkups performed during last pregnancy:

| Number of prenatal visits | Number of participants |
| --- | --- |
| 0 | 2 |
| 1-2 | 6 |
| 3-4 | 4 |
| 5-6 | 5 |
| 7-9 | 3 |

Two participants reported receiving no prenatal visits. Both lived in the Catatumbo area in the North of Santander in Colombia. One of them -a Venezuelan migrant- worked from 4 am to 7 pm. The other -a Colombian returnee- who lived in an area of social conflict. The latter mother gave birth at a relative’s home, which highlights how social violence in the region is also one of the causes of inaccessibility to health services and late diagnosis of syphilis.

The remaining 18 mothers had had between 1 and 9 prenatal visits, with Colombia being the place of attendance for the largest number of women (n=11); two had visits both in Colombia and Venezuela, and five had them in Venezuela. The main reason given by the mothers for not attending more check-ups was the economic cost of this medical appointment.

Of the total number of participants in the study, only two attended an adequate number of controls as per the Colombian standard of care, which recommends nine visits.

However, one of the two women who received prenatal visits was still only diagnosed with syphilis in her 39th week, preventing timely treatment.

The admission of these women to the hospital in Colombia for delivery and postnatal care provided them with immediate medical care and psycho-social support. It also allowed them to receive information together with the father of the child, and for both to be educated on the need for treatment for both the mother and her partner in order to eliminate syphilis.

> *The hospital gave me a second dose, the third one I have to go to Nueva EPS (health entity) for the other application, they told him to go to San Luis… the hospital was where I talked to him, I was coming out of the ICU and the only thing I was really worried about was my baby (P17, in her thirties)*.

### Theme: “The scarcity of support networks and machismo”

Participants broadly described limited social networks, as well as the relationships and tensions that are created within their families and with their sexual partners.

Only one of the participants, who is Colombian, was registered in the Colombian social protection program Families in Action, in which she received a monthly cash transfer of approximately US$100. Other participants did not report receiving economic support from the Colombian government. These participants relied on their partners, friends or relatives to support their economic needs, childcare and household chores. Participants commented on the role of grandmothers as caregivers in Venezuela, which enabled mothers to look for paid work in Colombia and to send back remittances for their children and for the rest of the family.

> *I got a job here in La Gabarra (municipality of the North of Santander, Colombia) and I started earning money and was able to send money to my siblings back in Venezuela, and they gave me food on the farm (P4, in her teens)*.

Nine participants commented on the importance of psychological counselling during diagnosis, hospital stay, and treatment of their newborns for congenital syphilis, considering it an important and necessary intervention for their mental health. This allowed them to better cope with their diagnosis, discuss it with their partner, and to support their partner’s willingness to adhere to treatment.

> *The syphilis thing stayed that way until I got out of the ICU, they talked to him and he started treatment (P17, in her thirties)*.

Regarding their partners, most women (15) reported that their relationships remained stable after their syphilis diagnosis. The remaining participants reported that their relationships had dissolved, in some cases maintaining communication only because of information about their newborns. This is due to the stressful and violent relationships they had before or after the news of the diagnosis of congenital syphilis. In one case, a woman had to continue to share accommodation with her former partner despite violent threats, due to economic necessity.

> *We live there in the house with him for the moment, but I have no contact with him, because we separated, because once while drinking with some friends they put things in his head, suddenly that the child was not his, and he in his anger was violent against me and was going to hit me with a knife in the belly (P17, in her thirties)*.

When asked about sexual practices, such as the use of condoms, two women reported that their partners had been reluctant to use them. However, the intervention of the health team, offering information to the couple, and especially to the man, helped reduce doubts and to maintain adherence to syphilis treatment and to prevent reinfection. The participants reported that sometimes the information had a positive impact, as follows:

> *It was a little difficult because he does not like (the use of condoms), the doctor told me it was for my health and, well, he understood (P10, in her thirties)*.

On the other hand, some women have had marital breakups due to the man’s refusal to use condoms because of discomfort and dissatisfaction.

> *He says that he does not like it, because I will not be with him again without a condom, that he does not like that, that it is ugly, he says (P10, in her thirties)*.

The mothers were also asked about reproductive autonomy, and whose responsibility it should be to prevent pregnancies when desired. Seven women said that it should be decided by the couple. Eleven mothers, especially those who had more than one child, emphasized that it should be the women, since they are responsible for the care of their children.

> *The woman, because the woman is the one who gets” screwed”, because all the responsibility that comes is on the woman (P17, in her thirties)*.

Likewise, when asked about whose responsibility it was to avoid a sexually transmitted infection, the majority of mothers (14), felt that both should prevent it, while others (3) alluded that it should be the woman.

> *It is not only the man who fails, but the woman as well; so if one has a home, one should protect it. If you are going to have something on the street, you should protect yourself, to protect your partner at home (P12, in her twenties)*.

A last group of participants felt it was the man’s responsibility, because the man is a “man”, and may be more likely to be unfaithful and promiscuous.

> *We slept together, in reality I took [the diagnosis of syphilis] neither very well nor very badly (), simply when I felt the change I let him go; the truth is he accepted it, the truth in front of me, I saw that he had someone else … he says it was her, moreover, I do not know if it was her or it was someone else (P13, in her twenties*.

Participants described feelings of fear of the partner’s reaction when he learns of the diagnosis; of what he will say or think about her because of it. In some cases, due to fear or a desire to avoid conflict, some participants reported that they did not disclose their diagnosis to their partners.

> *No, because he is going to believe that I am the one who infected him, he is not going to accept it (P15, in her twenties)*.
>
> *I have not told him either because men … they believe that it is one’s [own fault], that it is not their fault (P16, in her thirties)*.
>
> *Well, actually I told him that we had to have some injections, but I didn’t tell him what it was … because I was afraid (P10, in her thirties)*.

Men who already knew about the diagnosis, in some cases reportedly assumed attitudes of denial or blamed women for the diagnosis.

> *He didn’t take it in a very good way, because like all macho men … he blamed it all on me! (P14, in her twenties)*.

Participants reported that male partners were non-adherent to treatment due to fear of the pain that may be caused by administering the medication or because of work schedules that prevented them from going to a health center. In some cases, the woman refused sexual relations unless the man adhered to treatment, as in the following.

> *He didn’t want the injections because he is afraid, so I told him that if he didn’t get them, we would never be together again (P19, in her forties)*.

In this context, the health team, and in this case the physician and social worker, reportedly served as a mediator for the couples, ensuring that treatment was adopted as a means to avoid reinfection. However, the consequence of transmission to children was a motivation for some men to return for treatment.

> *He said what a shame, what a shame … he got angry, in the end he did not go through with the treatment … Now he says that he will go get treated, because of what happened to the girl (P20, in her thirties)*.

It is noteworthy that, despite the diagnosis, some participants reported continuing to have sexual relations without condoms, a practice that puts the couple at risk, since the treatment is still incomplete.

## DISCUSSION

The ongoing humanitarian crisis in Venezuela, and lack of access to basic necessities such as food or healthcare inside the country, has continued to produce an outflow of migration from the country, most acutely felt by bordering countries such as Colombia, which also has a history of displacement. The participants in our study - Venezuelan migrants and Colombian returnees- confirmed the dire conditions in their countries of origin or asylum, many of whom had crossed over to Colombia during their pregnancy due to lack of access to healthcare back in Venezuela.

The narratives of the mothers reveal the intersectionality of various conditions of vulnerability. They were young women, mostly migrants or Colombian returnees who have experienced forced displacement. Their social context is conditioned by low education and income, informal work often with long workdays, and few support networks, which are factors that prevail in populations that are in conditions of disadvantage and inequality [12, 26].

The compounded effects of lack of access to healthcare at critical life moments (eg. pregnancy) and the need to prioritize work, basic necessities, such as food and housing, and the needs of other family members, made it impossible for many women to access timely and regular prenatal care, resulting in undetected syphilis infections. These findings align with other findings that place the burden on family wellbeing on women who then often delay their own healthcare [27]. They also highlight that since the precarious socioeconomic conditions also act as a determinant that prevents access to health services.

Lack of reproductive health education and knowledge about syphilis as well as challenges related to intimate partnerships, complicated diagnosis, prevention, and treatment of syphilis. Blame from partners and resistance to treatment by partners highlight both the stigma of having a sexually transmitted infection (STI) and the power of masculinity (i.e., *machismo*), that prevented women from disclosing their diagnosis and encouraging partners to seek treatment. These situations can lead to reinfection and complications, since, if not diagnosed and treated in time, it produces irreversible long-term sequelae that facilitate transmission. All of which entails a greater risk of reinfection, as long as unprotected sexual relations are maintained.

## Limitations

This study included ten Colombian returnees; however, while their sociodemographic characteristics and reported experiences were similar to those of Venezuelan mothers, we may not have sufficiently achieved saturation that would identify unique experiences among this subpopulation. Further research with this group of women could be considered in order to expand the results in this special case.

## Conclusion

The findings show the vulnerable social position of the mothers of newborns with congenital syphilis, in contexts that can foster the reemergence of congenital syphilis. They reveal clear barriers to healthcare by many migrant and displaced women, in both Venezuela and in Colombia, providing evidence of the absence of effective national and international policies to reach these groups and prevent the spread of this syphilis [7, 18]. Addressing these barriers for migrants and returnees in Colombia may support Colombia’s achievement of goals set forth the WHO for the elimination of mother-to-child transmission [2].

The analysis above reveals the multiple missed opportunities for intervention via health policies, resulting in the outcomes of congenital syphilis. Proactive policies to provide prenatal healthcare to all pregnant women regardless of immigration status and documentation could be effective strategies for preventing mother-child transmission.

We especially recommend scaling up prenatal care access (inclusive of syphilis testing and treatment) in host communities with a high share of migrants and ensuring that refugees have access to this care.

Such efforts should be coupled with proactive strategies to reach male partners, provide psycho-social counselling, and to ensure adherence to treatment. Finally, the precarious conditions in which these women live highlight the need to create programs that provide material support for these families to address social determinants of health.

## Data Availability

Data sharing requests should be sent to the corresponding author and will be reviewed by lead investigators. Deidentified individual participant data and a data dictionary defining each field in the set are available upon request after a proposal is approved and the data use agreement is signed. The survey and consent form will be provided with study data upon approval of the data request and signed data use agreement.

## Acknowledgements

The project received financial support from the German Development Cooperation Agency, Deutsche Gesellschaft für Internationale Zusammenarbeit (GIZ) Bogotá 2.0 Initiative Project, administered by the Charitée under the umbrella COL-VAC-SIF project, and by the German Institute for Global and Area Studies. The project was implemented by CARE Colombia. We would like to thank the staff at the Hospital Universitario Erasmo Meoz as well as the CARE Colombia staff for their support in carrying out the interviews. Cole Simon assisted with the bibliography.

## Appendix

Interview guide (translated from Spanish)

**Guidelines for interviewing postpartum women with newborns with congenital syphilis at the Erasmo Meoz University Hospital (HUEM).**

**Informed consent________**.

**Interview number:________**

**Place of interview:________________**

**Date of interview:________________**

### Sociodemographic

1. Can you tell me a little about yourself: How old are you, what do you do for a living?
2. What is your marital status?
3. Can you tell me with whom you live and if you have any other children?
4. Can you tell me if you are currently living in Venezuela or Colombia, or if you go back and forth between the two countries?
5. Can you tell me what is your last year of school?
6. If you live in Colombia now, how long ago did you leave Venezuela? (Applicable only if you live in Colombia).
7. Can you tell me what documents do you have to enter or stay regularly in Colombia?
8. Can you tell me if you are affiliated to the Colombian health system, subsidized or contributory?
9. What is your relationship with the baby’s father?

### Socio-economic/contextual

1. For what reasons did you leave Venezuela or come to Colombia?
2. Can you tell me what are the main basic needs you need to solve?
3. During your pregnancy, have you decreased the portion size of the food you eat due to lack of money or other resources (access, availability, consumption and practices)?
4. Are you currently working at a job, what is your current job and what expenses are you able to cover?
5. Have you ever had to sleep on the street or in a place where you did not feel safe?
6. Do you have a support network to help you meet these needs?
7. Can you tell me what aspects you can improve in Colombia: food, education, employment, security?

### Knowledge of syphilis and access to protection and treatment measures

1. What are the most important sources of sex education for you?

School, parents, siblings, friends, internet, church, other?

1. What do you know about syphilis?
2. How and when did you learn that you have syphilis?
3. What did you know about syphilis before your diagnosis? Methods of protection, how can you get it?
4. Have you had prenatal checkups in this pregnancy? If yes, how many?

How many?

In Colombia, in Venezuela, or in both countries?

Can you tell me if you were tested for syphilis during your pregnancy? Do you remember how many weeks pregnant you were?

If positive, were you offered treatment?

If you were offered treatment, what were the obstacles to accessing or completing treatment? [ask for details] How many doses of penicillin did you receive?

How many doses of penicillin did you receive?

5. Where you were tested for syphilis, were you offered free treatment for syphilis immediately?
6. Can you tell me what the health professional told you about syphilis treatment for yourself and what he/she told you for your partner?”

Possible next questions:

7. Where you were tested for syphilis, did they explain to you the importance of treating your partner?
8. Where you were tested for syphilis, did they explain to you the importance of getting treated for syphilis every week?
9. Where you were tested for syphilis, was the importance of using a condom during the week of treatment explained to you?
10. When you were told you had syphilis, were you told to go to another health care setting, given a shot, or given an injection?
11. Did you tell your partner that you had syphilis and for that reason, did you tell your partner that he also needed treatment?
12. If you talked to your partner, what motivated you, what enabled you to talk openly with your partner (e.g., relationship factors, any services or information offered, being aware of treatment for the partner) Emphasize this aspect.
13. If you did not tell your partner what were the reasons?
14. During the weeks of treatment did you use condoms for all sexual intercourse, if not, can you tell me the reasons?
15. Did your partner understand the importance of curing syphilis and receive the full treatment (three injections), if not, what were the reasons?

### Questions about access to health care in general

1. What is your experience in accessing (emphasize) health care to prevent or cure sexually transmitted infections or to prevent or control pregnancy in Venezuela?
2. What is your experience with the same in Colombia?
3. If you had difficulties in accessing health services, what were the obstacles?

### Relationships

Can you tell me about your relationship with your partner or the father of the baby?

[Consider asking about where you live, your immigration status (whether you live in Colombia, pendular or in transit), whether you live together and/or have other children together).

1. How would you characterize your relationship with your partner or the father of the baby?
2. If you live with the baby’s father:

Can you tell me if your partner shares other responsibilities of offices in the home?

3. Can you tell me if you can negotiate with the father/your partner about condom use?
4. Do you have difficulty using methods (condoms) to protect against syphilis (with your partner)?
5. Who do you think should avoid pregnancy, the man or the woman? [Response categories: Woman’s, mostly the woman’s, both, mostly the man’s, mostly the man’s] In agreeing on this, what aspects do you take into account?

For example, if it is influenced by the fact that one of the two works and the other is at home or by family customs.

6. Who do you think should avoid a sexually transmitted infection, the man or the woman? To agree on this, what aspects do you take into account?

For example, does the fact that one of you works and the other is at home or family customs play a role?

7. How old were you at the time of your first sexual intercourse?
8. In the last year, how many sexual partners have you had?
9. Have you ever had unwanted sexual contact?
10. How many pregnancies have you had? How old were you when you had your first pregnancy?
11. When you have sex, do you ever, sometimes, usually, always use a condom?

### Finally, do you have any other comments you would like to share?

Do you have any other comments you would like to share with us?

## Reference List

1. Tao YT, Gao TY, Li HY, Ma YT, Li HJ, Xian-Yu CY, et al. Global, regional, and national trends of syphilis from 1990 to 2019: the 2019 global burden of disease study. BMC Public Health. 2023 Apr 24;23(1):754. doi: 10.1186/s12889-023-15510-4.

2. World Health Organization, Pan American Health Organization. Orientaciones mundiales sobre los criterios y procesos para la validación de la eliminación de la transmission maternoinfantil del VIH, la sífilis y el virus de la hepatitis B. [Global guidance on criteria and processes for validation: Elimination of mother- to-child transmission of HIV, syphilis and hepatitis B virus]. 2021. Available from: https://iris.paho.org/bitstream/handle/10665.2/56219/9789275325858_spa.pdf?sequence=1&isAllowed=y. Spanish.

3. Arando Lasagabaster M, Otero Guerra L. Syphilis. Enferm Infecc Microbiol Clin (Engl Ed). 2019 Jun-Jul;37(6):398–404. doi: 10.1016/j.eimc.2018.12.009. Epub 2019 Feb 7.

4. Camacho-Montano AM, Nino-Alba, R, Paez-Castellanos E. Congenital syphilis with hydrops fetalis: report of four cases in a general referral hospital in Bogota, Colombia between 2016- 2020. Rev Colomb Obstet Ginecol. 2021 Jun 30;72(2):149–161. doi: 10.18597/rcog.3591.

5. Sankaran D, Partridge E, Lakshminrusimha S. Congenital Syphilis-An Illustrative Review. Children (Basel). 2023 Jul 29;10(8):1310. doi: 10.3390/children10081310.

6. Lim J, Yoon SJ, Shin JE, Han JH, Lee SM, Eun HS, et al. Outcomes of infants born to pregnant women with syphilis: a nationwide study in Korea. BMC Pediatr. 2021 Jan 22;21(1):47. doi: 10.1186/s12887-021-02502-9.

7. Keuning MW, Kamp GA, Schonenberg-Meinema D, Dorigo-Zetsma JW, van Zuiden JM, Pajkrt D. Congenital syphilis, the great imitator—case report and review. Lancet Infect Dis. 2020 Jul;20(7):e173–e179. doi: 10.1016/S1473-3099(20)30268-1. Epub 2020 Jun 2.

8. Departamento Nacional de Planeacion; Ministerio de Salud y Proteccion Social. Comportamiento de la sífilis gestacional y la sífilis congénita en Colombia, semanas epidemiológicas 01 a 39 de 2017 a 2023. [Behavior of gestational syphilis and congenital syphilis in Colombia, epidemiological weeks 01 to 39 from 2017 to 2023]. 2023. Available from: https://www.ins.gov.co/buscador-eventos/BoletinEpidemiologico/2023_Bolet%C3%ADn_epidemiologico_semana_41.pdf. Spanish.

9. Perez C. Comportamiento de la vigilancia en salud pública de la sífilis gestacional y congénita en Colombia, con énfasis en población migrante. [Behavior of public health surveillance of gestational and congenital syphilis in Colombia, with emphasis on the migrant population]. Subcluster SSR. 2024 Feb: 38. Spanish.

10. Regional Inter-agency Coordination Platform. R4V Latin America and the Caribbean, Venezuelan Refugees and Migrants in the Region - Nov 2023 [Internet]. 2023 Nov 30. Available from: https://www.r4v.info/en/document/r4v-latin-america-and-caribbean-venezuelan-refugees-and-migrants-region-nov-2023.

11. Colombia’s Refugee Crisis and Integration Approach Explained. USA for UNHCR. 2024 Apr 18. Available from: https://www.unrefugees.org/news/colombia-s-refugee-crisis-and-integration-approach-explained/#:~:text=By%20the%20end%20of%202022,all%20internal%20displacements%20since%202004.

12. Adaleti R, Kansak N, Aslan M, Balkose G, Toptan H, Dincer SD, et al. Comparison of syphilis seropositivity between non-immigrant and immigrant populations in the Anatolian side of Istanbul, Turkiye: Results of five-years retrospective study. North Clin Istanb. 2022 Dec 22;9(6):590–594. doi: 10.14744/nci.2021.80688.

13. Stevenson M, Guillen J, Ortiz J, Ramirez Correa JF, Page KR, Barriga Talero MA, et al. Syphilis prevalence and correlates of infection among Venezuelan refugees and migrants in Colombia: findings of a cross-sectional biobehavioral survey. Lancet Reg Health Am. 2024 Jan 16:30:100669. doi: 10.1016/j.lana.2023.100669.

14. Cuellar Segura C, Rodriguez Rodriguez A, Roa Sanchez A, Daza Huerfano J, Marin Rodriguez D. Comportamiento de la sífilis gestacional y congénita en población migrante venezolana atendida en los servicios de salud en Colombia 2020, 2021. [Behavior of gestational and congenital syphilis in the Venezuelan migrant population treated in health services in Colombia, 2020-2021]. Bogotá; Ministerio de Salud y Protección Social. Available from: https://www.minsalud.gov.co/sites/rid/Lists/BibliotecaDigital/RIDE/VS/ED/GCFI/nota-politica-n1-comportamiento-sifilis-gestacional-congenita-migrantes-2020-2021.pdf. Spanish.

15. Brizuela V, Bahamondes L, Gomez Ponce de Leon R, Aslanyan G, Feletto M, Bonet M, et al. Fortalecimiento de la investigacion local para abordar la salud y los derechos sexuales y reproductivos de las personas migrantes de Venezuela y América Central. [Strengthening local research to address the sexual and reproductive health and rights of migrants from Venezuela and Central America]. Rev Panam Salud Publica. 2023 Mar 3:47:e37. doi: 10.26633/RPSP.2023.37. Spanish.

16. Consejo Danes para Refugiados. Monitoreo de protección: Colombia, México, Perú y Venezuela 2022. [Protection monitoring: Colombia, Mexico, Peru and Venezuela Annual Report 2022] [Internet]. Maracaibo: Consejo Danes para Refugiados. 2022. Available from: https://reliefweb.int/attachments/91ff4f11-0e74-493b-86ad-81cb25747f67/Informe_Regional_MdP_2022_VF.pdf. Spanish.

17. Angeleri S, Ramirez Bolivar L, Arroyave Velasquez L. Derecho a la atención en salud para las personas migrantes en situación irregular en Colombia: Entre estándares normativos y barreras prácticas. [Right to health care for migrants in an irregular situation in Colombia: Between regulatory standards and practical barriers]. Bogotá: Dejusticia; 2024 Mar 20. Available from: https://www.dejusticia.org/publication/derecho-a-la-atencion-en-salud-para-las-personas-migrantes-en-situacion-irregular-en-colombia-entre-estandares-normativos-y-barreras-practicas/. Spanish.

18. Ministerio de Salud y Protección Social; United Nations Populations Fund. Guía de práctica clínica (GPC) basada en la evidencia para la atención integral de la sífilis gestacional y congénita. [Evidence-based Clinical Practice Guide (CPG) for the comprehensive care of gestational and congenital syphilis]. 2014. Available from: https://www.minsalud.gov.co/sites/rid/Lists/BibliotecaDigital/RIDE/INEC/IETS/gpc-guia-corta-sifilis.pdf. Spanish.

19. Ministerio de Salud y Protección Social. [Resolution Number 3280] [Internet]. Dec 2018. https://www.minsalud.gov.co/sites/rid/Lists/BibliotecaDigital/RIDE/DE/DIJ/resoluci on-3280-de-2018.pdf.

20. Husserl E. Experience and Judgment: Investigations in a Genealogy of Logic. 1st Span ed. Mexico City: Universidad Nacional Autónoma de Mexico; 1980.

21. Schutz A, Luckmann T. The Structures of the Life-World. 1st Span ed. Buenos Aires: Amorrortu; 2004.

22. Lincoln YS, Guba EG. Naturalistic Inquiry. Beverly Hills (CA): SAGE Publications; 1985.

23. Tong A, Sainsbury P, Craig J. Consolidated criteria for reporting qualitative research (COREQ): a 32-item checklist for interviews and focus groups. Int J Qual Health Care. 2007 Dec;19(6):349–57. doi: 10.1093/intqhc/mzm042. Epub 2007 Sep 14.

24. Glaser BG, Strauss AL. The Discovery of Grounded Theory: Strategies for Qualitative Research. New Brunswick (NJ): AldineTransaction; 1967.

25. Strauss A, Corbin J. Basics of qualitative research. Techniques and procedures for developing grounded theory. 1st Span ed. Medellin (CO): Universidad de Antioquia; 2002.

26. Fajardo AO, Eslava-Schmalbach J. Desigualdades en la incidencia de sífilis congénita relacionada en la condiciones de vida, Bogotá Colombia 2013-2014. [Inequalities in the incidence of congenital syphilis related to living conditions, Bogota Colombia 2013-2014]. Cad saude colet. 2020 Dec;28(4): 510–517. doi: 10.1590/1414-462x202028040441. Spanish.

27. Valenzuela ME, Lucia Scuro M, Vaca-Trigo L. Desigualdad, crisis de los cuidados y migración del trabajo doméstico remunerado en América Latina. [Inequality, the care crisis and migration of paid domestic work in Latin America ECLAC]. 2020 Dec. Available from: https://www.cepal.org/es/publicaciones/46537-desigualdad-crisis-cuidados-migracion-trabajo-domestico-remunerado-america. Spanish.

